# COVID-19-related school closures, United States, July 27, 2020 – June 30, 2022

**DOI:** 10.1101/2023.08.31.23294738

**Authors:** Nicole Zviedrite, Ferdous Jahan, Sarah Moreland, Faruque Ahmed, Amra Uzicanin

## Abstract

As part of a multi-year project that monitored illness-related school closures, we conducted systematic daily online searches from July 27, 2020–June 30, 2022, to identify public announcements of COVID-19-related school closures (COVID-SCs) in the US lasting ≥1 day. We explored the temporospatial patterns of COVID-SCs and analyzed associations between COVID-SCs and national COVID-19 surveillance data. COVID-SCs reflected national surveillance data: correlation was highest between COVID-SCs and both new PCR test positivity (correlation coefficient, r = 0·73, CI: 0·56, 0·84) and new cases (r = 0·72, CI: 0·54, 0·83) in school year (SY) 2020-21, and with hospitalization rates among all ages (rs = 0·81, CI: [0·67, 0·89]) in SY 2021-22. The number of reactive COVID-SCs during SYs 2020-21 and 2021-22 greatly exceeded previously observed numbers of illness-related reactive school closures in the US, notably being nearly 5-fold greater than reactive closures observed during the 2009 H1N1 Pandemic (H1N1pdm09 virus).

**Article summary line:** COVID-19-related school closures occurred annually in the US and their temporal patterns mirror the general patterns of COVID-19 activity at the national level as observed through routine COVID-19 epidemiological surveillance.

## Introduction

While unplanned school closures occur every year, outside of a pandemic only a small minority (∼1%) are associated with infectious disease while the majority are due to weather and natural disasters (1). However, the initial months (February-June 2020) of the coronavirus disease 2019 (COVID-19) pandemic in the United States (US) led to unprecedented, nearly-simultaneous, nationwide implementation of kindergarten through twelfth grade (K-12) school closures throughout the US, as a part of a wider effort to slow virus transmission and reduce disease burden (2). In most communities, these early pandemic-related closures were implemented pre-emptively as a non-pharmaceutical intervention before community transmission was high. In contrast, the subsequent COVID-19-related school closures occurring in the 2020–21 and 2021–22 school years (SYs) were predominantly reactive, i.e., occurring after infection had affected students and/or staff.

As school re-convened for SYs 2020–21 and 2021–22, schools and districts were faced with the challenge of providing in-person education and services during the ongoing pandemic. In a previous analysis, we described the transition to online learning that occurred between February-June 2020, following the COVID-19 related preemptive closures of schools and school districts (2). The subsequent two pandemic-affected school years (2020-21 and 2021-22) were characterized by the deployment of various education modalities, including education that was fully in-person, fully distance learning, or a hybrid model (3, 4, 5). In both years, among schools offering in-person learning (fully or hybrid), school closures continued to be implemented in response to local transmission dynamics and policies, as well as due to other consequences of the pandemic (vaccination of staff and students and side effects of vaccination, teacher and staff shortages, and pandemic related mental health issues).

This study describes trends in reactive COVID-19-related K-12 school closures (COVID-SCs) during the COVID-19 pandemic in the US from July 27, 2020, to June 30, 2022 and analyzes associations between COVID-SCs and national level COVID-19 epidemiological surveillance data.

## Methods

### Data collection

We conducted daily systematic online searches from July 27, 2020 through June 30, 2022, to identify public announcements of unplanned, illness-related K-12 school closures in the United States. Searches were conducted in Google and Google News using the following terms: “school closed” and either “COVID,” “COVID-19,” or “coronavirus.” Additionally, the search string “(academy OR school OR district OR class) AND (close OR closing OR closure OR cancel OR cancelled) AND (coronavirus OR corona OR ‘COVID-19’ OR COVID OR ‘novel coronavirus’)” was employed in a Google Alert. Publicly available COVID-19-related school closure dashboards identified during these routine searches were also checked, including those published online by school districts, state and local education authorities, and private entities. All school closure announcements were saved as portable document files (PDFs) prior to data abstraction. Only announcements mentioning COVID-19 as a reason for closure were included for analysis. Additional details on the search strategy and data abstraction processes are available in previously published methodology (1, 2). Data were not collected from July 1 – July 25, 2021 as this coincided with school summer break in the US. Fully in-person and hybrid learning modalities were classified as open, and modalities without in-person learning (i.e., fully distanced learning and closed) were classified as closed. Unplanned closure was defined as a transition from being open to being closed for in-person instruction, for ≥1 day. If a school/district reopened for ≥1 day and then closed again, the subsequent closure was counted as a new occurrence of closure. For closures that spanned both unplanned and planned closure days, such as those contiguous with weekends or planned holidays, only unplanned closure days were counted. For closures for which a reopening date could not be identified following closure, length of closure was assumed to be one day. Schools that delayed in-person learning at the start of the school year, through closure or full distance-learning, were not captured in this data collection. For each identified COVID-SC, we abstracted school or school district name, state, dates of closure and reopening, as well as reasons for closure as reported in the announcements.

### Contextualizing school closures

To better understand the characteristics of schools and districts experiencing COVID-SCs, each closure event was matched to publicly available data downloaded from the National Center for Education Statistics (NCES) using the respective NCES district or school ID (identified using district/school name and location) (6, 7). Some districts and schools experienced multiple COVID-SC events during the study period and, therefore, appear more than once in the resulting dataset. For each public school and public school district, COVID-SC data were matched with the respective year (2020-21, 2021-22) of NCES data, and for private schools, COVID-SCs were matched with the most recent year of data available (2019-2020) in NCES. NCES data include information such as the number of schools per district, the number of students and staff, the number of students eligible in the federal free or reduced-price school lunch programs (only for public schools and public school districts), urbanicity, and grade span data. We excluded: schools with pre-kindergarten or transitional kindergarten students only; school districts with no schools; schools with no students; vocational, special education, and alternative schools with missing values for student enrollment; and permanent distance-learning-only or adult education schools.

### Epidemiologic data

We gathered publicly available COVID-19 surveillance data from July 27, 2020 – June 30, 2022, including daily new cases and death counts (8), weekly hospitalization rates (9), and daily PCR positivity (10) for the duration of the study period. Hospitalization rates were reported per epidemiological week and corresponding weekly averages were calculated for new cases, deaths, and PCR positivity at the national level. Epidemiological weeks run Sunday – Saturday, with epidemiological week one being the first week to hold four or more days from the new calendar year.

### Data analysis

We described characteristics of school closures according to the data abstracted from public announcements. Specific reasons for COVID-SCs were summarized by grouping them into 12 non-mutually exclusive categories under two primary themes (1) transmission-related reasons (COVID-19 cases, suspected cases, increased student absenteeism, increased staff absenteeism, cluster or widespread transmission in the community, state or local guidance/mandate to close schools in response to COVID-19, to clean/disinfect school facilities, and other reasons related to COVID-19 mitigation including testing, contact tracing, quarantine of students and staff, prevention of holiday-related surge, death of staff member, critical lack of community resources – including contact tracers, and noncompliance with Governor’s executive orders) and (2) non-transmission-related reasons (COVID-19 vaccinations, teacher/staff shortages, for student/staff mental health, and other reasons associated with COVID-19 including staff protests of in-person learning, protests over mask policies, transportation issues specific to COVID-19, lack of resources specific to COVID-19, and in order to work on the COVID-19 mitigation plan).

We compared weekly patterns of COVID-SCs and with COVID-19 epidemiological surveillance data at the national level (new cases and deaths, hospitalization rates, and lab test positivity). We calculated Spearman rank correlations (r) and 95% confidence intervals (CI) to evaluate these relationships (at alpha=0.05) during the school years. The final week of each calendar year was excluded from analysis, as this week coincides with school winter break in the US. P-value calculation for the Spearman’s rank correlation was based on Fisher’s Z-transformation. Analysis was conducted using SAS 9.4 (SAS Institute Inc., Cary, North Carolina). Visualizations were created using Microsoft Excel and Microsoft Power BI. Results for each school year are presented sequentially.

Additional analyses, including calculations of in-person school days lost, the frequency and patterns of repeat closures of schools for COVID-19, the cumulative incidence of COVID-SCs by states, and bivariate and multivariable regression analyses, are described in supplementary material.

## Results

### School Year 2020–2021

From July 27, 2020, to June 30, 2021, an estimated 19,273 COVID-SCs (Table 1) were experienced by 16,890 unique schools. About three-quarters closed as part of district-wide closures. More than 11 million students were affected, losing more than a combined 159 million in-person student-days. The majority of COVID-SCs were observed in the first half of the school year (Aug – Dec 2020) and they peaked in epidemiological week 47, the week prior to the US Thanksgiving holiday (Fig 1). The median number of in-person schools days lost per closure nationally was 10 days (interquartile range (IQR): 3-23) (Fig S1), however it reached to over 20 days in seven states (California, Colorado, Illinois, Indiana, Kentucky, Minnesota, and Nevada) (Fig S2). Most schools experiencing COVID-SCs, experienced only one COVID-SC during the school year (Table S1). However, 12·1% (n= 2,036) experienced between two and seven COVID-SCs during the school year, with two being the most common (n=1,756, 86·2%).

**Figure 1.**
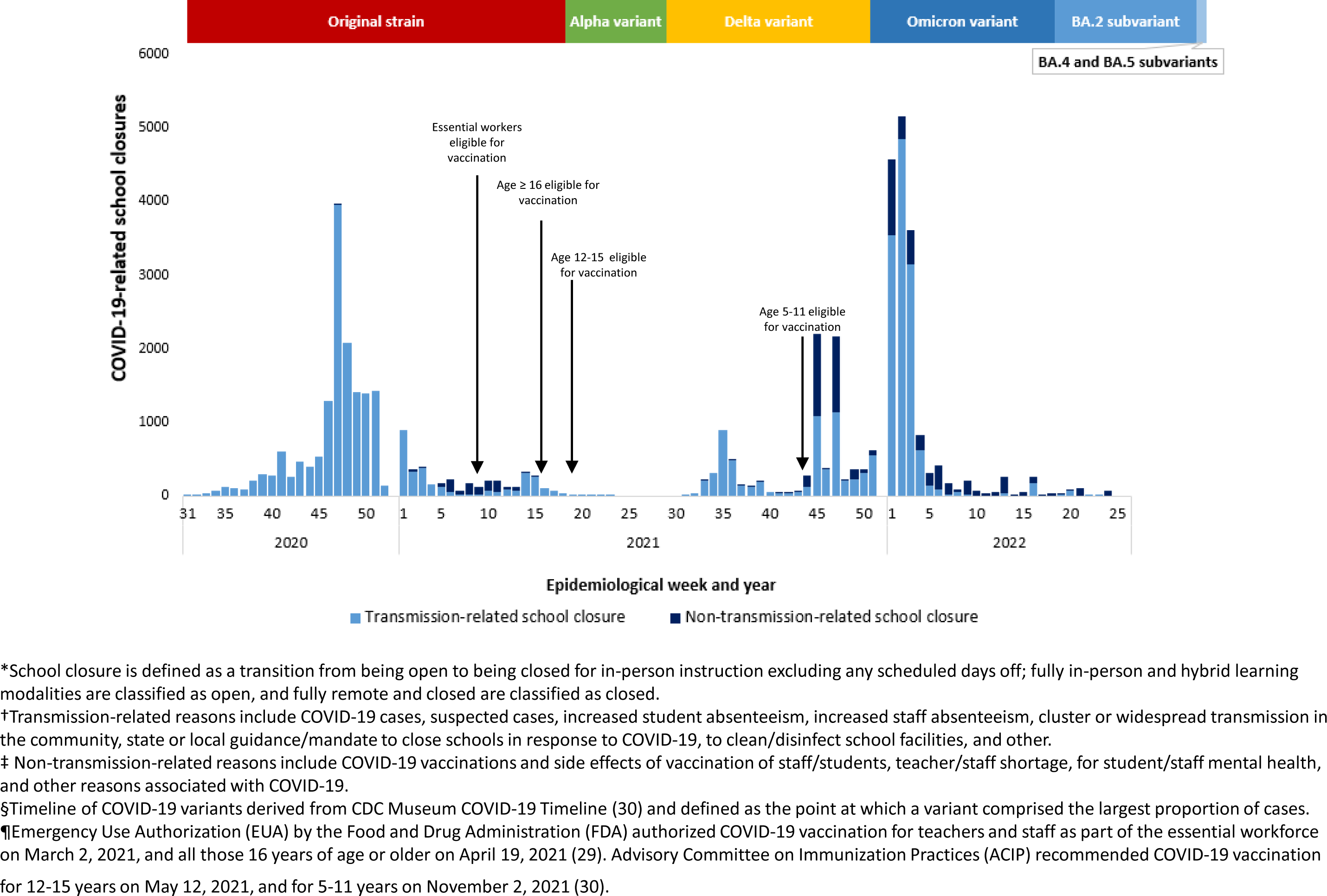
COVID-19-related School Closures^*†‡^ and Dominant COVID-19 Variants^§^, United States, July 27, 2020 – June 30, 2022.

**Table 1.**
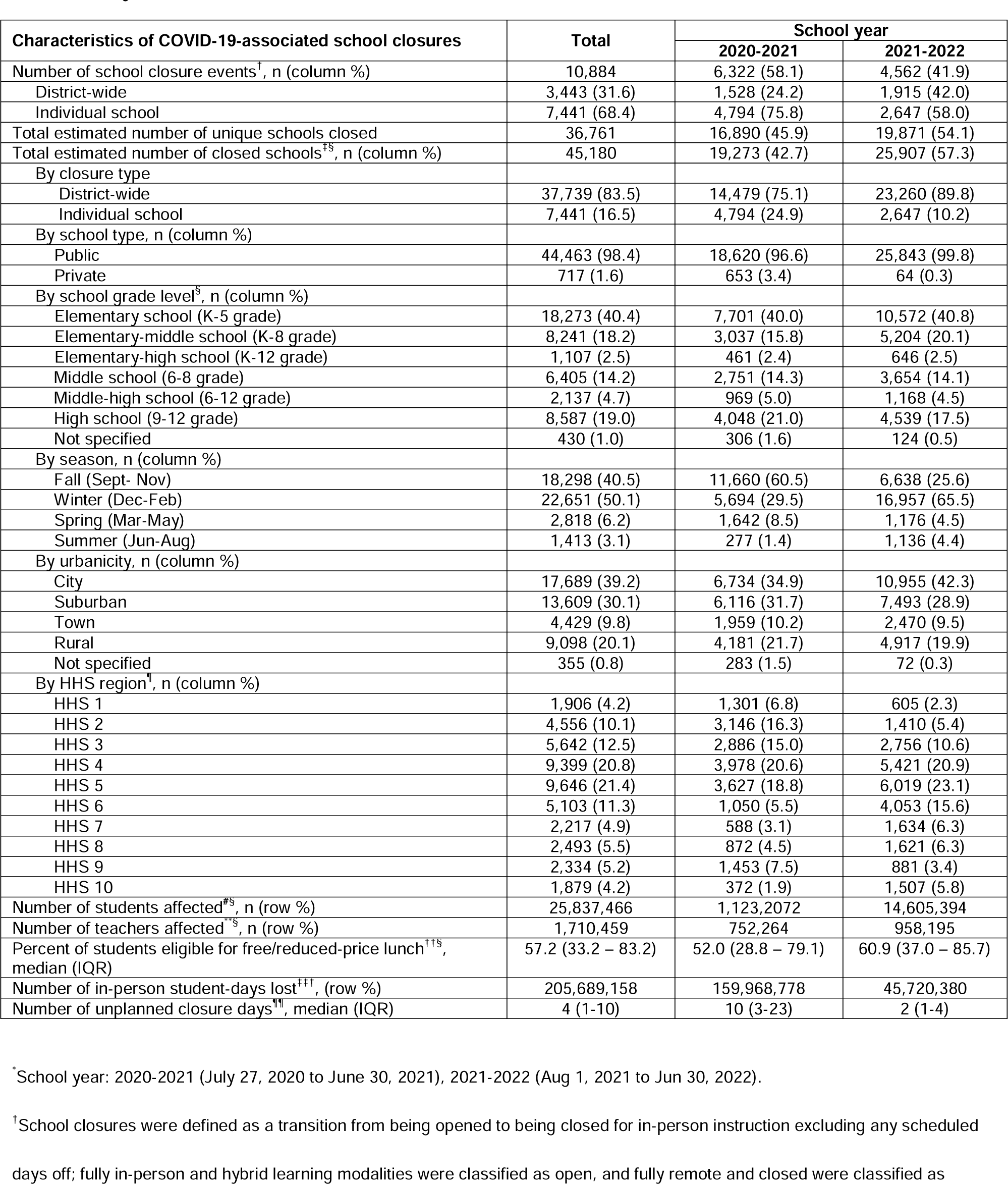
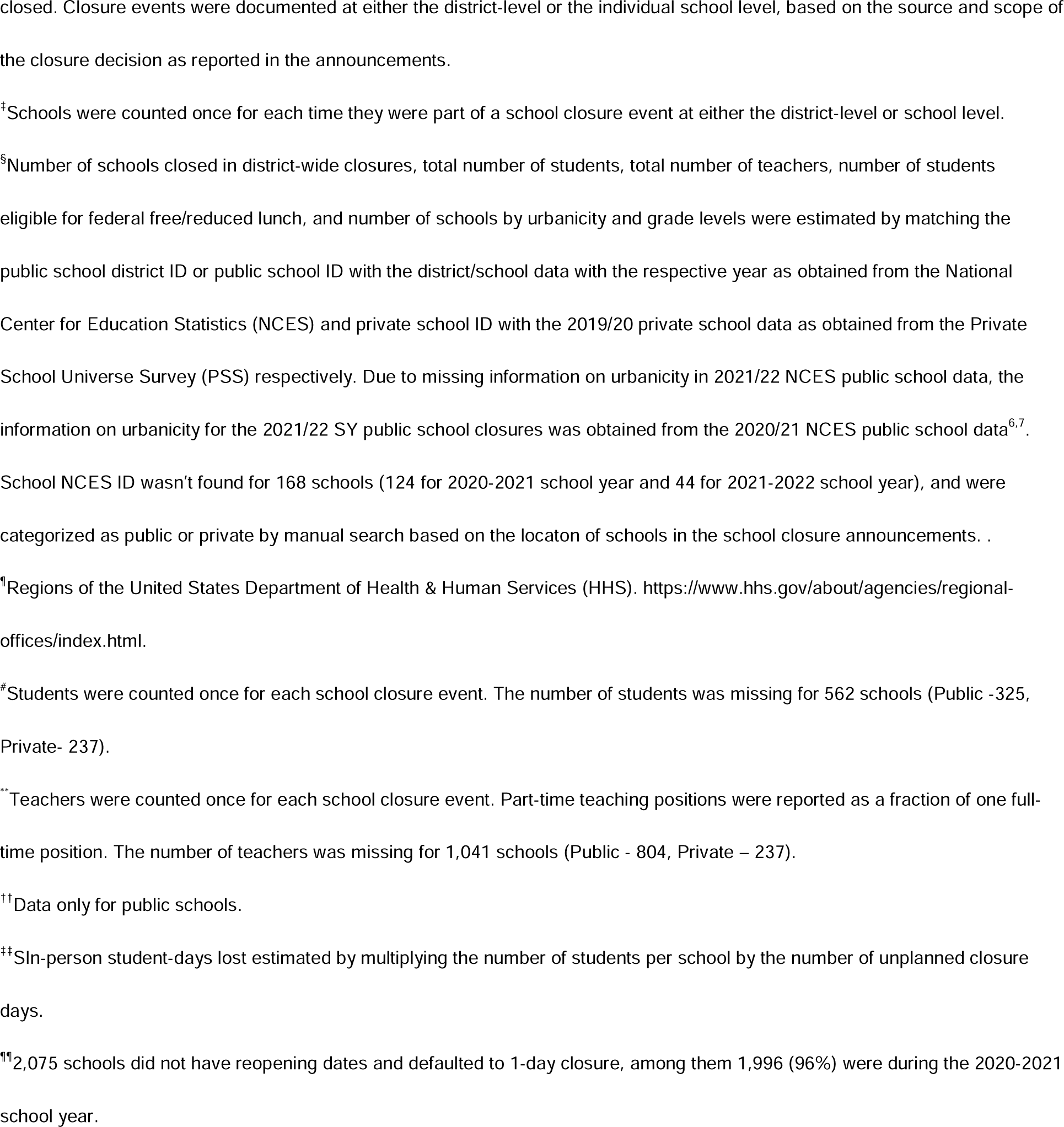
Characteristics of COVID-19-associated school closures by school year* – United States, July 27, 2020 – June 30, 2022.

Among the announcements reporting reasons for closure, the most common reason was having positive cases in the school, followed by clusters or widespread transmission in the community (Table 2). Additionally, state and local mandates to close schools in response to COVID-19 accounted for more than 20% of closures. Mandates were issued either as part of an ongoing policy triggered by reaching a covid threshold, such as the positivity rate of testing (11), and second, in response to local surges in cases (12, 13). While few COVID-SCs were attributed to non-transmission related reasons such as vaccination of staff and students or non-specific teacher shortages attributed to the pandemic, nearly 30% of COVID-SCs between epidemiological weeks 6 and 11 of 2021 were due to vaccination of staff and side effects of vaccination (Fig 1).

**Table 2.** Reasons* for COVID-19 Related K-12 School Closure events– United States, July 27, 2020—June 30, 2022^†^.

| Reasons for school closure decision stated in announcement of COVID-19-related closure events<sup>‡</sup> | Total |  | 2020-2021 |  | 2021-2022 |  |
| --- | --- | --- | --- | --- | --- | --- |
|  | District | School | District | School | District | School |
| Number of COVID-19 related closure events | 3,443 | 7,441 | 1,528 | 4,794 | 1,915 | 2,647 |
| School closure announcement mentions only COVID-19 | 85 (2.5) | 123 (1.7) | 43 (2.8) | 76 (1.6) | 42 (2.2) | 49 (1.9) |
| School closure announcement mentions COVID-19 and specific reasons | 3,358 (97.5) | 7,318(98.3) | 1,485 (97.2) | 4,718 (98.4) | 1,873 (97.8) | 2,598 (98.1) |
| Transmission-related reasons |  |  |  |  |  |  |
| Due to positive case(s) | 1,845 (53.6) | 4,511 (60.6) | 724 (47.4) | 2,829 (59.0) | 1,121 (58.5) | 1,682 (61.5) |
| In student(s) | 758 (22.0) | 1,412 (19.0) | 233 (15.2) | 651 (13.6) | 525 (27.4) | 761 (28.7) |
| In staff member(s) | 763 (22.2) | 1,359 (18.3) | 226 (14.8) | 575 (12.0) | 537 (28.0) | 784 (29.6) |
| In household member of student/staff | 15 (0.4) | 24 (0.3) | 11 (0.7) | 12 (0.3) | 4 (0.2) | 12 (0.5) |
| In visitor(s) | 2 (0.1) | 0 (0.0) | 2 (0.1) | 0 (0.0) | 0 (0.0) | 0 (0.0) |
| Due to suspected case(s) | 78 (2.3) | 224 (3.0) | 64 (4.2) | 173 (3.6) | 14 (0.7) | 51 (1.9) |
| In student(s) | 23 (0.7) | 70 (0.9) | 21 (1.4) | 48 (1.0) | 2 (0.1) | 22 (0.8) |
| In staff member(s) | 33 (1.0) | 71 (1.0) | 27 (1.8) | 53 (1.1) | 6 (0.3) | 18 (0.7) |
| In household member of student/staff | 0 (0.0) | 5 (0.1) | 0 (0.0) | 5 (0.1) | 0 (0.0) | 0 (0.0) |
| Due to increased student absenteeism | 515 (15.0) | 732 (9.8) | 56 (3.7) | 132 (2.8) | 459 (24.0) | 600 (31.3) |
| Due to increased staff absenteeism | 1,022 (29.7) | 1,955 (26.3) | 183 (12.0) | 536 (11.2) | 839 (43.8) | 1,419 (53.6) |
| Due to cluster or widespread transmission in the community | 1,096 (31.8) | 1,918 (25.8) | 727 (47.8) | 1,556 (32.5) | 369 (19.3) | 362 (13.7) |
| Due to state or local guidance/mandate | 348 (10.1) | 1,146 (15.4) | 318 (20.8) | 1,091 (22.8) | 30 (1.6) | 55 (2.1) |
| To clean/disinfect classrooms, buildings, and facilities | 240 (7.0) | 682 (9.2) | 102 (6.7) | 562 (11.7) | 138 (7.2) | 120 (4.5) |
| Other<sup>§</sup> | 356 (10.3) | 750 (10.1) | 146 (9.6) | 532 (11.1) | 210 (11.0) | 218 (8.2) |
| Non-transmission-related reasons |  |  |  |  |  |  |
| Vaccination of staff/students | 83 (2.4) | 56 (0.8) | 66 (4.3) | 49 (1.0) | 17 (0.9) | 7 (0.3) |
| Side effects of vaccination | 9 (0.3) | 18 (0.2) | 9 (0.6) | 18 (0.4) | 0 (0.0) | 0 (0.0) |
| Teacher shortage | 135 (3.9) | 158 (2.1) | 20 (1.3) | 26 (0.5) | 115 (6.0) | 132 (5.0) |
| Mental health | 158 (4.6) | 48 (0.6) | 0 (0.0) | 0 (0.0) | 158 (8.3) | 48 (1.8) |
| Other<sup>¶</sup> | 17 (0.5) | 32 (0.4) | 0 (0.0) | 10 (0.2) | 17 (0.9) | 22(0.8) |

<sup>‡</sup>Reasons are recorded as stated in the school closure announcements.
<sup>†</sup>School year: 2020-2021 (July 27, 2020 to June 30, 2021), 2021-2022 (Aug 1, 2021 to Jun 30, 2022).
<sup>‡</sup>Categories are not mutually exclusive because a closure announcement may attribute the closure to more than one factor and/or there may be more than one announcement that contributes the closure to different factors.
<sup>§</sup>Other reasons include contact tracing, quarantine of students and staff, prevention of holiday-related surge, precautionary measure as a concern of community spread due to union wanting work stoppage, death of staff member, unable to find substitute teachers, transportation issues, critical lack of community resources – including contact tracers, testing, out of an abundance of caution, and flu/other respiratory and/or enteric virus-related illnesses, internet outage, facility issues, and noncompliance with Governor's executive orders.
<sup>¶</sup>Other reasons include staff protesting in-person learning, protest over mask policy, transportation issue, lack of resources, allowing time for testing, and work on the COVID-19 mitigation plan.

Transmission-related COVID-SCs were strongly correlated with weekly COVID-19 testing positivity rates (r=0·73, CI: 0·56, 0·84) and with new COVID-19 cases (r=0·72, CI: 0·54, 0·83) (Table 3). Transmission-related COVID-SCs were moderately correlated with both new COVID-19 deaths by week (r=0·51, CI: 0·25, 0·69) and weekly laboratory confirmed COVID-19-associated hospitalization rates for all ages (r=0·64, CI: 0·42, 0.78) (Table 4). Age-specific correlation with hospitalization rates varied, with the 5-17 years age group, which aligns with the K-12 student population, having the weakest correlation (r=0·37, CI: 0·09, 0·60) and correlations strengthening for each subsequent older age group (Table 4). The peak in weekly COVID-SCs preceded the peaks in COVID-19 disease surveillance indicators (new cases, deaths, and percent positive PCR tests) by roughly 6-8 weeks (Fig 2.a, Fig 3.a).

**Figure 2.**
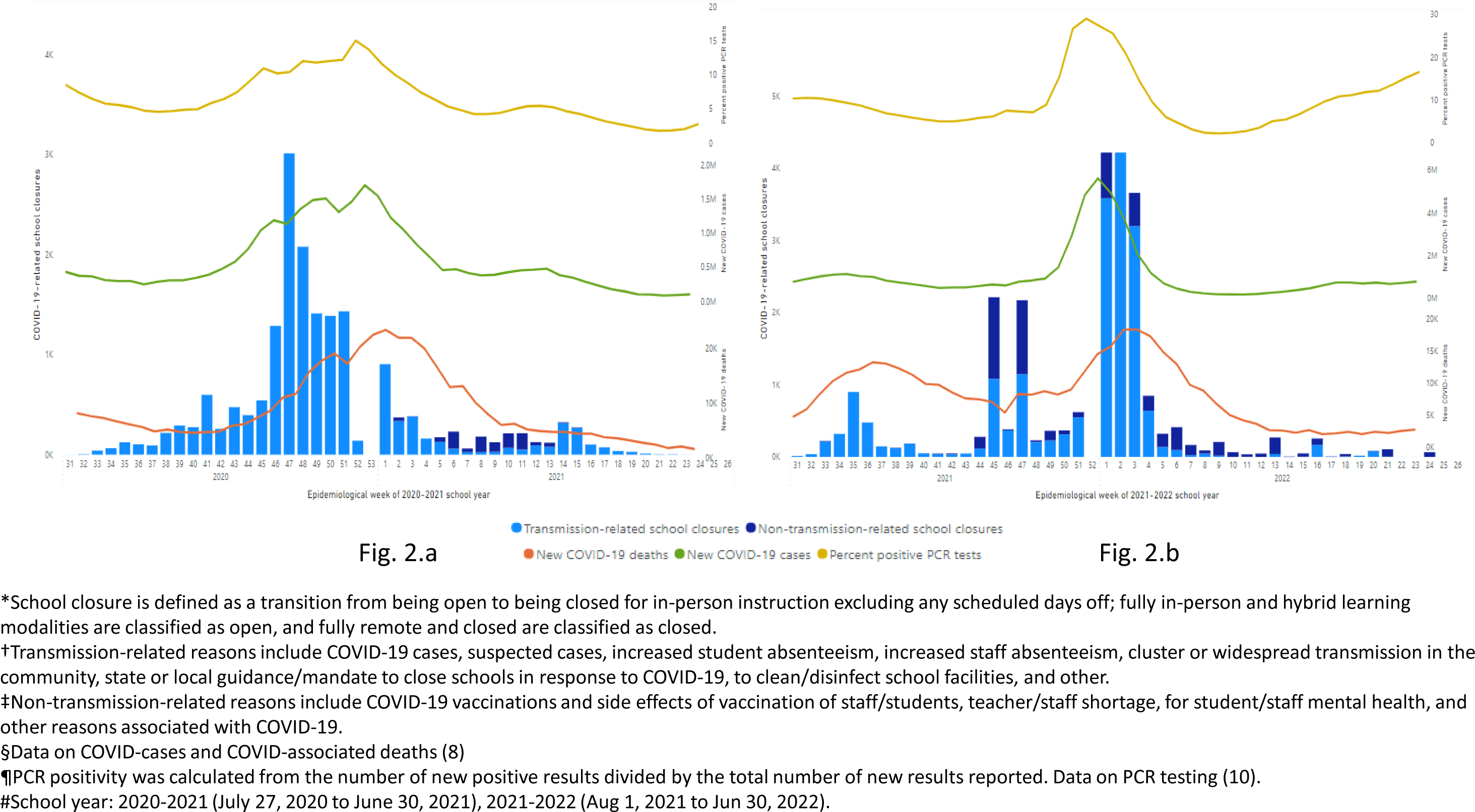
COVID-19-related School Closures^*†‡^ and COVID-related cases^§^, deaths^§^, and PCR positivity^¶^ by school year^#^, United States, July 27, 2020 – June 30, 2022.

**Figure 3.**
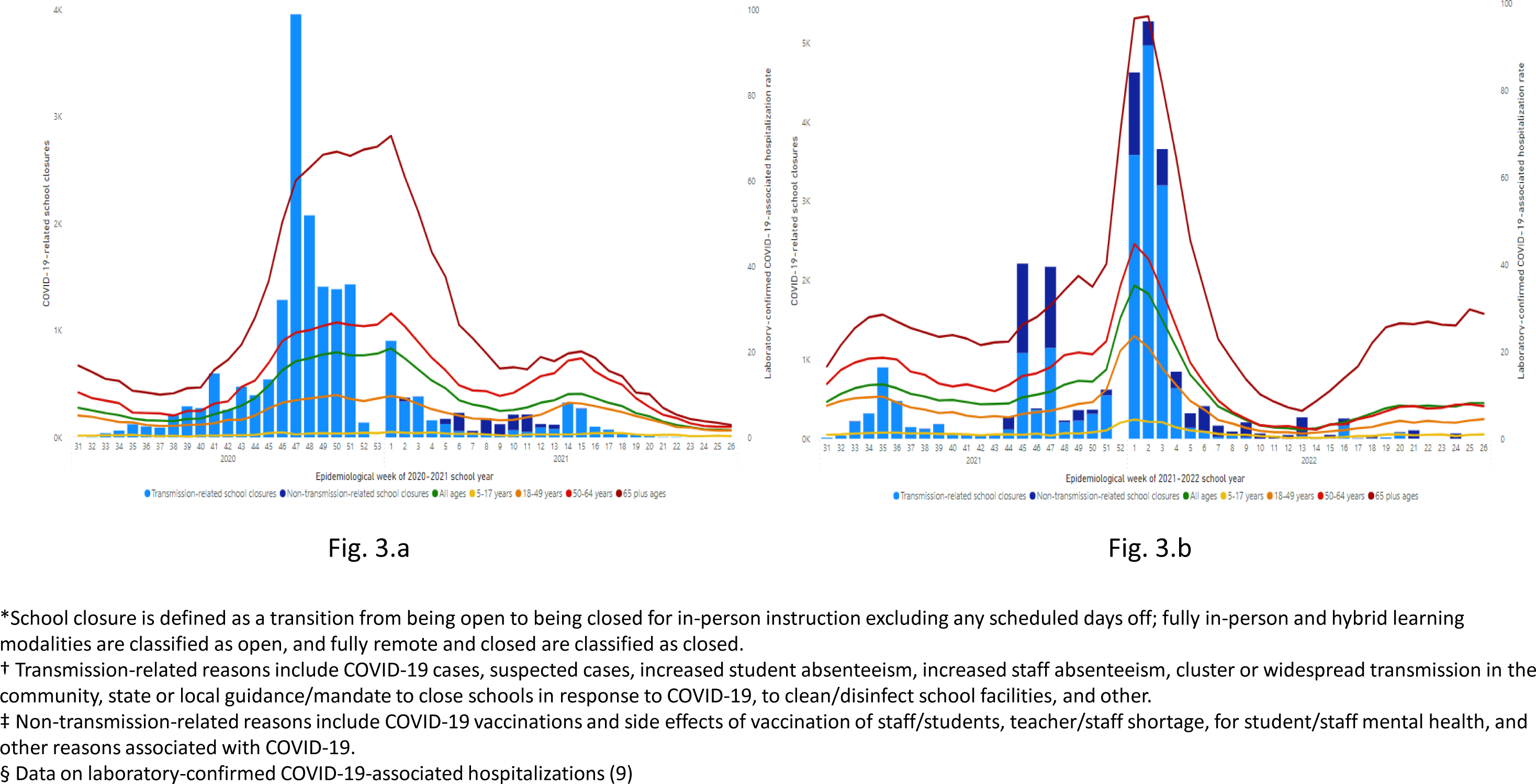
COVID-19-related School Closures^*†‡^ and Laboratory-confirmed COVID-19-related Hospitalization by Age^§^, United States, July 27, 2020 – June 30, 2022.

**Table 3.**
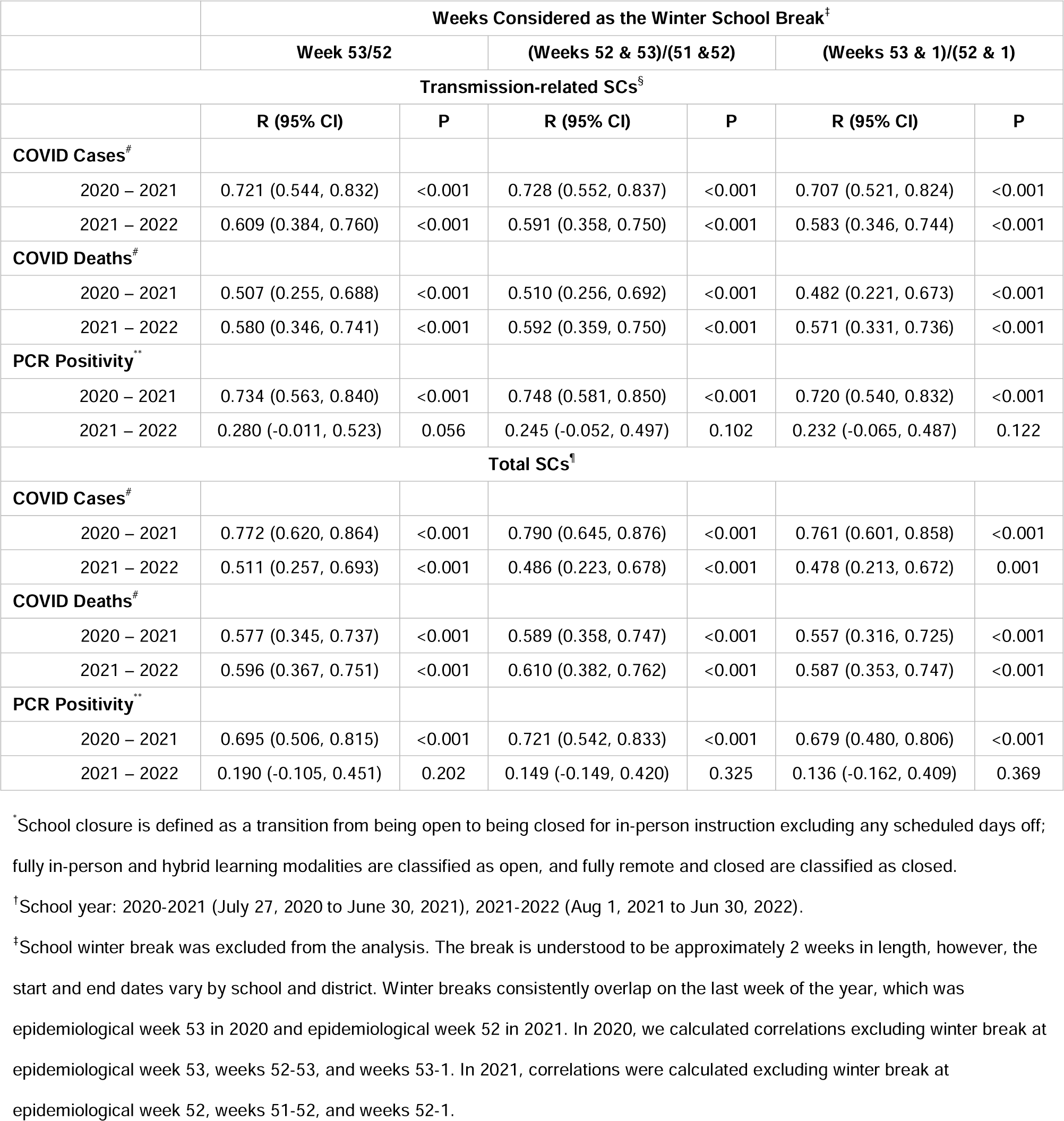
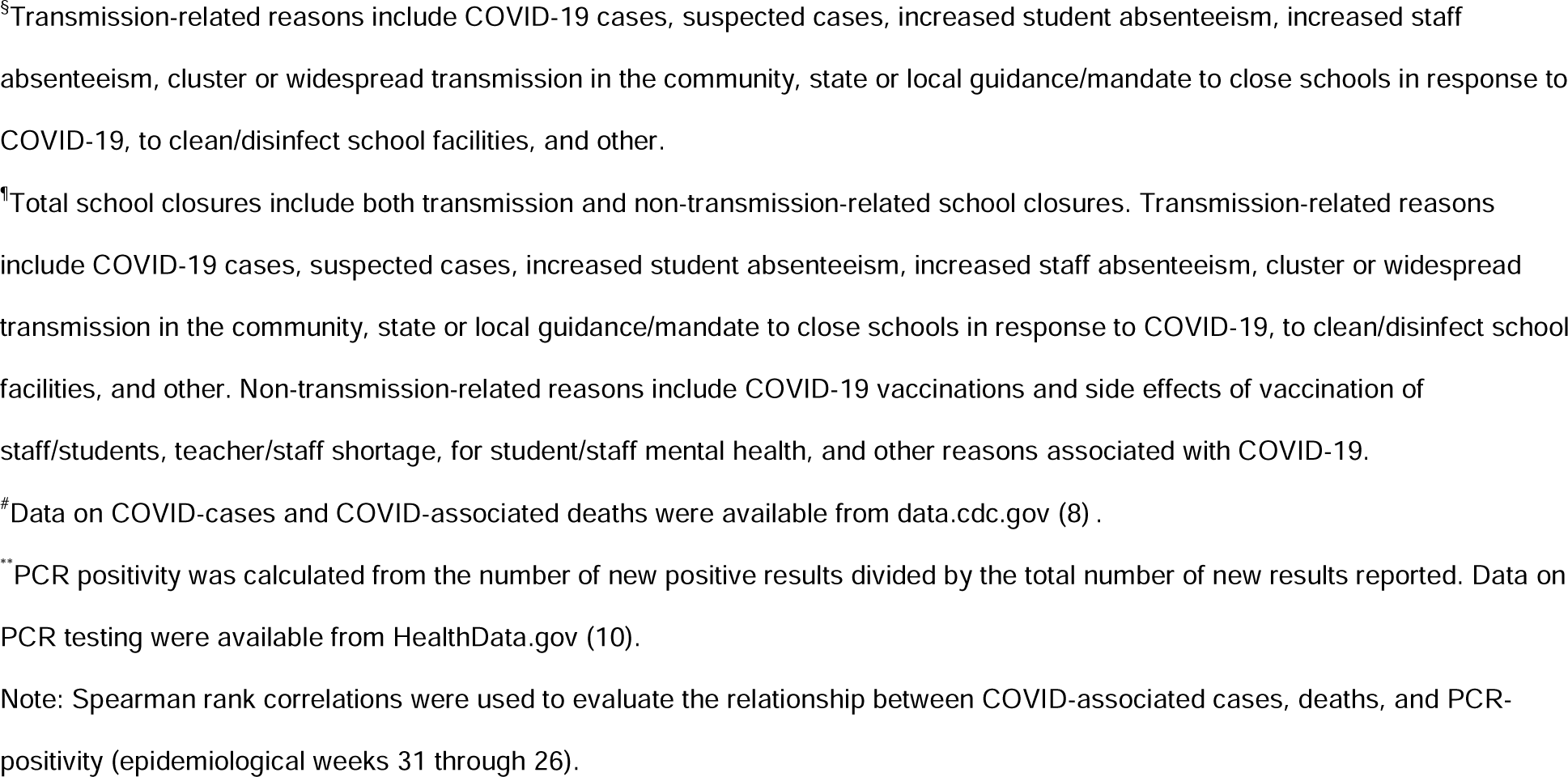
Weekly correlation between COVID-related school closures (SC)* and COVID-related cases, deaths, and PCR positivity by school year^†^– United States, July 27, 2020— June 30, 2022.

|  | Weeks Considered as the Winter School Break<sup>‡</sup> |  |  |  |  |  |
| --- | --- | --- | --- | --- | --- | --- |
|  | Week 53/52 |  | (Weeks 52 & 53)/(51 & 52) |  | (Weeks 53 & 1)/(52 & 1) |  |
| Transmission-related SCs<sup>§</sup> |  |  |  |  |  |  |
|  | R (95% CI) | P | R (95% CI) | P | R (95% CI) | P |
| **COVID Cases<sup>#</sup>** |  |  |  |  |  |  |
| 2020 – 2021 | 0.721 (0.544, 0.832) | <0.001 | 0.728 (0.552, 0.837) | <0.001 | 0.707 (0.521, 0.824) | <0.001 |
| 2021 – 2022 | 0.609 (0.384, 0.760) | <0.001 | 0.591 (0.358, 0.750) | <0.001 | 0.583 (0.346, 0.744) | <0.001 |
| **COVID Deaths<sup>#</sup>** |  |  |  |  |  |  |
| 2020 – 2021 | 0.507 (0.255, 0.688) | <0.001 | 0.510 (0.256, 0.692) | <0.001 | 0.482 (0.221, 0.673) | <0.001 |
| 2021 – 2022 | 0.580 (0.346, 0.741) | <0.001 | 0.592 (0.359, 0.750) | <0.001 | 0.571 (0.331, 0.736) | <0.001 |
| **PCR Positivity<sup>\*\*</sup>** |  |  |  |  |  |  |
| 2020 – 2021 | 0.734 (0.563, 0.840) | <0.001 | 0.748 (0.581, 0.850) | <0.001 | 0.720 (0.540, 0.832) | <0.001 |
| 2021 – 2022 | 0.280 (-0.011, 0.523) | 0.056 | 0.245 (-0.052, 0.497) | 0.102 | 0.232 (-0.065, 0.487) | 0.122 |
| **Total SCs<sup>¶</sup>** |  |  |  |  |  |  |
| **COVID Cases<sup>#</sup>** |  |  |  |  |  |  |
| 2020 – 2021 | 0.772 (0.620, 0.864) | <0.001 | 0.790 (0.645, 0.876) | <0.001 | 0.761 (0.601, 0.858) | <0.001 |
| 2021 – 2022 | 0.511 (0.257, 0.693) | <0.001 | 0.486 (0.223, 0.678) | <0.001 | 0.478 (0.213, 0.672) | 0.001 |
| **COVID Deaths<sup>#</sup>** |  |  |  |  |  |  |
| 2020 – 2021 | 0.577 (0.345, 0.737) | <0.001 | 0.589 (0.358, 0.747) | <0.001 | 0.557 (0.316, 0.725) | <0.001 |
| 2021 – 2022 | 0.596 (0.367, 0.751) | <0.001 | 0.610 (0.382, 0.762) | <0.001 | 0.587 (0.353, 0.747) | <0.001 |
| **PCR Positivity<sup>\*\*</sup>** |  |  |  |  |  |  |
| 2020 – 2021 | 0.695 (0.506, 0.815) | <0.001 | 0.721 (0.542, 0.833) | <0.001 | 0.679 (0.480, 0.806) | <0.001 |
| 2021 – 2022 | 0.190 (-0.105, 0.451) | 0.202 | 0.149 (-0.149, 0.420) | 0.325 | 0.136 (-0.162, 0.409) | 0.369 |

\*School closure is defined as a transition from being open to being closed for in-person instruction excluding any scheduled days off; fully in-person and hybrid learning modalities are classified as open, and fully remote and closed are classified as closed.
<sup>†</sup>School year: 2020-2021 (July 27, 2020 to June 30, 2021), 2021-2022 (Aug 1, 2021 to Jun 30, 2022).
<sup>‡</sup>School winter break was excluded from the analysis. The break is understood to be approximately 2 weeks in length, however, the start and end dates vary by school and district. Winter breaks consistently overlap on the last week of the year, which was epidemiological week 53 in 2020 and epidemiological week 52 in 2021. In 2020, we calculated correlations excluding winter break at epidemiological week 53, weeks 52-53, and weeks 53-1. In 2021, correlations were calculated excluding winter break at epidemiological week 52, weeks 51-52, and weeks 52-1.
<sup>§</sup>Transmission-related reasons include COVID-19 cases, suspected cases, increased student absenteeism, increased staff absenteeism, cluster or widespread transmission in the community, state or local guidance/mandate to close schools in response to COVID-19, to clean/disinfect school facilities, and other.
<sup>¶</sup>Total school closures include both transmission and non-transmission-related school closures. Transmission-related reasons include COVID-19 cases, suspected cases, increased student absenteeism, increased staff absenteeism, cluster or widespread transmission in the community, state or local guidance/mandate to close schools in response to COVID-19, to clean/disinfect school facilities, and other. Non-transmission-related reasons include COVID-19 vaccinations and side effects of vaccination of staff/students, teacher/staff shortage, for student/staff mental health, and other reasons associated with COVID-19.
<sup>#</sup>Data on COVID-cases and COVID-associated deaths were available from data.cdc.gov (8).
<sup>\*\*</sup>PCR positivity was calculated from the number of new positive results divided by the total number of new results reported. Data on PCR testing were available from HealthData.gov (10).
Note: Spearman rank correlations were used to evaluate the relationship between COVID-associated cases, deaths, and PCR-positivity (epidemiological weeks 31 through 26).

**Table 4.**
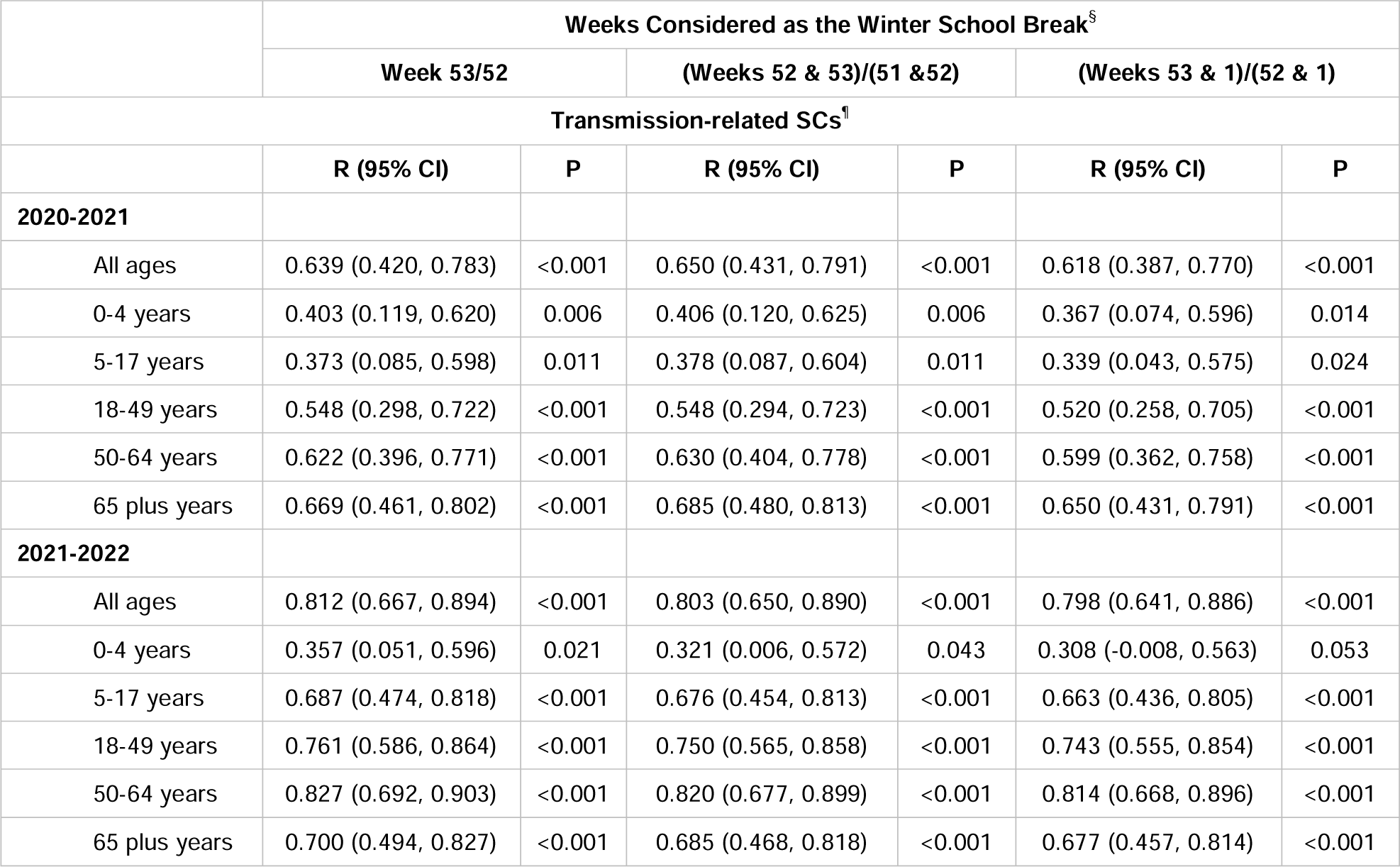
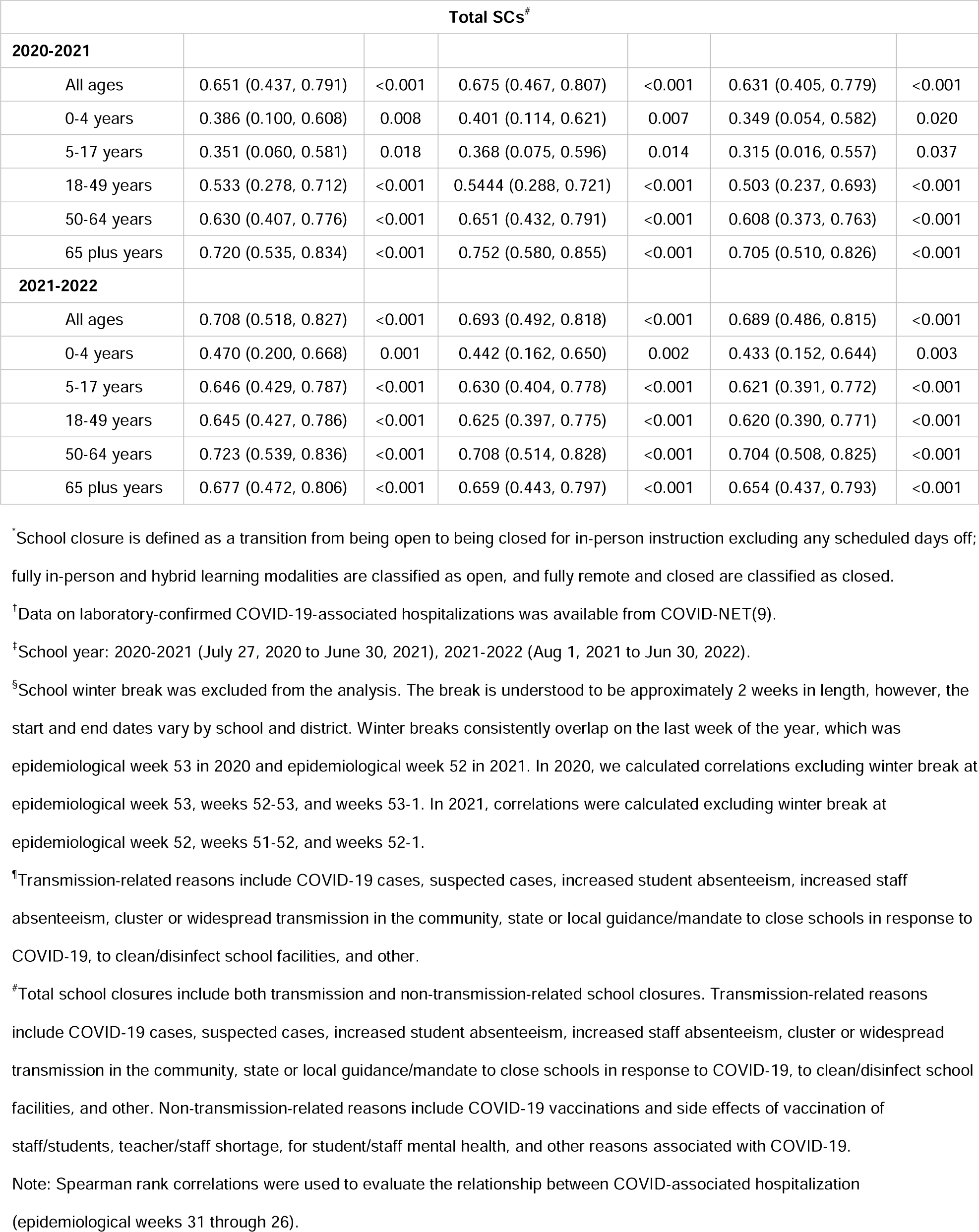
Weekly correlation between COVID-related school closures (SC)* and laboratory-confirmed COVID-19-associated hospitalizations^†^ by school year^‡^– United States, July 27, 2020—June 30, 2022.

|  | Weeks Considered as the Winter School Break<sup>§</sup> |  |  |  |  |  |
| --- | --- | --- | --- | --- | --- | --- |
|  | Week 53/52 |  | (Weeks 52 & 53)/(51 & 52) |  | (Weeks 53 & 1)/(52 & 1) |  |
| Transmission-related SCs<sup>†</sup> |  |  |  |  |  |  |
|  | R (95% CI) | P | R (95% CI) | P | R (95% CI) | P |
| 2020-2021 |  |  |  |  |  |  |
| All ages | 0.639 (0.420, 0.783) | <0.001 | 0.650 (0.431, 0.791) | <0.001 | 0.618 (0.387, 0.770) | <0.001 |
| 0-4 years | 0.403 (0.119, 0.620) | 0.006 | 0.406 (0.120, 0.625) | 0.006 | 0.367 (0.074, 0.596) | 0.014 |
| 5-17 years | 0.373 (0.085, 0.598) | 0.011 | 0.378 (0.087, 0.604) | 0.011 | 0.339 (0.043, 0.575) | 0.024 |
| 18-49 years | 0.548 (0.298, 0.722) | <0.001 | 0.548 (0.294, 0.723) | <0.001 | 0.520 (0.258, 0.705) | <0.001 |
| 50-64 years | 0.622 (0.396, 0.771) | <0.001 | 0.630 (0.404, 0.778) | <0.001 | 0.599 (0.362, 0.758) | <0.001 |
| 65 plus years | 0.669 (0.461, 0.802) | <0.001 | 0.685 (0.480, 0.813) | <0.001 | 0.650 (0.431, 0.791) | <0.001 |
| 2021-2022 |  |  |  |  |  |  |
| All ages | 0.812 (0.667, 0.894) | <0.001 | 0.803 (0.650, 0.890) | <0.001 | 0.798 (0.641, 0.886) | <0.001 |
| 0-4 years | 0.357 (0.051, 0.596) | 0.021 | 0.321 (0.006, 0.572) | 0.043 | 0.308 (-0.008, 0.563) | 0.053 |
| 5-17 years | 0.687 (0.474, 0.818) | <0.001 | 0.676 (0.454, 0.813) | <0.001 | 0.663 (0.436, 0.805) | <0.001 |
| 18-49 years | 0.761 (0.586, 0.864) | <0.001 | 0.750 (0.565, 0.858) | <0.001 | 0.743 (0.555, 0.854) | <0.001 |
| 50-64 years | 0.827 (0.692, 0.903) | <0.001 | 0.820 (0.677, 0.899) | <0.001 | 0.814 (0.668, 0.896) | <0.001 |
| 65 plus years | 0.700 (0.494, 0.827) | <0.001 | 0.685 (0.468, 0.818) | <0.001 | 0.677 (0.457, 0.814) | <0.001 |

| Total SCs<sup>#</sup> |  |  |  |  |  |  |
| --- | --- | --- | --- | --- | --- | --- |
| **2020-2021** |  |  |  |  |  |  |
| All ages | 0.651 (0.437, 0.791) | <0.001 | 0.675 (0.467, 0.807) | <0.001 | 0.631 (0.405, 0.779) | <0.001 |
| 0-4 years | 0.386 (0.100, 0.608) | 0.008 | 0.401 (0.114, 0.621) | 0.007 | 0.349 (0.054, 0.582) | 0.020 |
| 5-17 years | 0.351 (0.060, 0.581) | 0.018 | 0.368 (0.075, 0.596) | 0.014 | 0.315 (0.016, 0.557) | 0.037 |
| 18-49 years | 0.533 (0.278, 0.712) | <0.001 | 0.5444 (0.288, 0.721) | <0.001 | 0.503 (0.237, 0.693) | <0.001 |
| 50-64 years | 0.630 (0.407, 0.776) | <0.001 | 0.651 (0.432, 0.791) | <0.001 | 0.608 (0.373, 0.763) | <0.001 |
| 65 plus years | 0.720 (0.535, 0.834) | <0.001 | 0.752 (0.580, 0.855) | <0.001 | 0.705 (0.510, 0.826) | <0.001 |
| **2021-2022** |  |  |  |  |  |  |
| All ages | 0.708 (0.518, 0.827) | <0.001 | 0.693 (0.492, 0.818) | <0.001 | 0.689 (0.486, 0.815) | <0.001 |
| 0-4 years | 0.470 (0.200, 0.668) | 0.001 | 0.442 (0.162, 0.650) | 0.002 | 0.433 (0.152, 0.644) | 0.003 |
| 5-17 years | 0.646 (0.429, 0.787) | <0.001 | 0.630 (0.404, 0.778) | <0.001 | 0.621 (0.391, 0.772) | <0.001 |
| 18-49 years | 0.645 (0.427, 0.786) | <0.001 | 0.625 (0.397, 0.775) | <0.001 | 0.620 (0.390, 0.771) | <0.001 |
| 50-64 years | 0.723 (0.539, 0.836) | <0.001 | 0.708 (0.514, 0.828) | <0.001 | 0.704 (0.508, 0.825) | <0.001 |
| 65 plus years | 0.677 (0.472, 0.806) | <0.001 | 0.659 (0.443, 0.797) | <0.001 | 0.654 (0.437, 0.793) | <0.001 |

\*School closure is defined as a transition from being open to being closed for in-person instruction excluding any scheduled days off; fully in-person and hybrid learning modalities are classified as open, and fully remote and closed are classified as closed.
†Data on laboratory-confirmed COVID-19-associated hospitalizations was available from COVID-NET(9).
‡School year: 2020-2021 (July 27, 2020 to June 30, 2021), 2021-2022 (Aug 1, 2021 to Jun 30, 2022).
§School winter break was excluded from the analysis. The break is understood to be approximately 2 weeks in length, however, the start and end dates vary by school and district. Winter breaks consistently overlap on the last week of the year, which was epidemiological week 53 in 2020 and epidemiological week 52 in 2021. In 2020, we calculated correlations excluding winter break at epidemiological week 53, weeks 52-53, and weeks 53-1. In 2021, correlations were calculated excluding winter break at epidemiological week 52, weeks 51-52, and weeks 52-1.
¶Transmission-related reasons include COVID-19 cases, suspected cases, increased student absenteeism, increased staff absenteeism, cluster or widespread transmission in the community, state or local guidance/mandate to close schools in response to COVID-19, to clean/disinfect school facilities, and other.

### School Year 2021–2022

From August 1, 2021, to June 30, 2022, more than 14·6 million students were affected by an estimated 25,907 COVID-SCs (Table 1). Among unique schools which experienced COVID-SCs, the majority closed once (77·5%), while more than 20% closed 2-8 times (Table S1). Most closures occurred in the first three weeks of 2022, peaking at epidemiological week 2 (Fig 1). The median number of in-person school days lost per closure was 2 days (IQR: 1-4) (Fig S1).

Closures occurred in all 50 states and DC, with >1,000 closures in each of seven states (Texas, Illinois, Missouri, Ohio, North Carolina, Georgia, and Tennessee) (Fig S3.a) accounting for more than a third of all COVID-SCs observed during the school year. More than half of the schools in Alabama, Nevada, and Oregon (51·6%, 51·3, and 51·2%, respectively) and between 40% and 50% of schools in eight additional states (Oklahoma, Maryland, Tennessee, Utah, Kentucky, Colorado, Virginia, and Nebraska; Fig S3.b) experienced a COVID-SC.

Most district and school level COVID-SCs were attributed to positive cases in the schools (58·5% and 61·5%, respectively) (Table 2). Non-transmission-related reasons accounted for a greater number of COVID-SCs than in the prior school year, particularly those due to long-term teacher/staff shortages (shortages related to hiring and retention challenges, rather than directly linked to current disease transmission).

Transmission-related COVID-SCs were strongly correlated with weekly hospitalization rates for all ages (r=0·81, CI: 0·67, 0·89), among which correlation was moderate for the 5-17 year old age group (r=0·69, CI: 0·47, 0·82) and strong for the three adult aged groups (Table 4). Transmission-related COVID-SCs were moderately correlated with new COVID-19 cases (r=0·61, CI: 0·38, 0·76) and new COVID-19 deaths (r=0·58 CI: 0·35, 0·74), while there was no significant correlation for percent positive COVID-19 PCR tests (Table 3). The peak in weekly COVID-SCs occurred within 1-2 weeks of the peaks in weekly new COVID-19 cases, percent PCR positivity, and hospitalization rates, and preceded the peak in weekly new COVID-19 deaths by 3 weeks (Fig 2.b, Fig 3.b).

## Discussion

Our study describes COVID-19 school closures as school systems and communities grappled with ongoing disease transmission during a rapidly evolving pandemic. The COVID-SCs we analyzed reflect the SARS-CoV-2 spread among school-aged children and staff, as demonstrated by the correlation between COVID-SCs and COVID-19 epidemiologic surveillance. The large increase in illness-related closures during SYs 2020-21 and 2021-22, which was nearly 5-fold higher than those observed during severe influenza seasons, including the 2009 H1N1 Pandemic (H1N1pdm09 virus) (14) and subsequent moderate and severe influenza seasons (namely 2017-18, 2018-19, and 2019-20), was nearly fully attributed to COVID-19 (15). This increase likely reflected both ongoing transmission of SARS-CoV-2 and the greater clinical severity of COVID-19 infection among children as compared to influenza, as demonstrated by both higher rates of hospitalization and higher rates of ICU admission among those 18 and younger (16, 17).

The most frequently documented reason for closure in both SYs was COVID-19 cases in the school or school district. School year 2020-21 also had a significant number of closures due to increased community transmission and due to state/local mandates to close schools in response to COVID-19, consistent with the changing incidence of COVID-19 among both the general population and children over the study period (8, 9). COVID-SCs due to teacher absenteeism (due to illness of themselves or others) and teacher/staff shortages were more common in SY 2021-22 than in SY 2020-21. While teacher shortages were also reported prior to the pandemic, they were exacerbated by pandemic (18, 19).

In SY 2020-21, the number of schools open to in-person education varied throughout the semester with some schools teaching in-person, some relying solely on distance learning, some utilizing hybrid or mixed methods, and others moving between different modalities in response to disease transmission and related guidance throughout the school year (5). COVID-19 transmission during the 2020-21 SY was primarily dominated by the ancestral strain of SARS-CoV2, which would only later be supplanted by new variants as the predominating virus in circulation (20, 21). The majority of COVID-SCs in SY 2020-21 occurred in the first semester (August – December 2020), with the bulk occurring in the two weeks leading up to the Thanksgiving holiday (November 26, 2020) break. Despite recommendations from the CDC against travel during the holiday (22), travel during Thanksgiving week of 2020 (November 22-28, 2020) was at its highest since the start of the pandemic eight months prior (23, 24). During this time of increased social gatherings and movement, many schools that closed prior to Thanksgiving chose to stay shuttered until after the subsequent winter break (25, 26, 27). Annual (by school year) peaks among the epidemiological surveillance data occurred in January 2021. Following Emergency Use Authorization (EUA) by the Food and Drug Administration (FDA) of the first COVID-19 vaccine in December 2020 (28), teachers and staff became eligible for vaccination as part of the essential workforce on March 2, 2021 and all those 16 years of age or older on April 19, 2021 (29). Subsequently, the succeeding Alpha variant began to predominate in early April 2021 (21, 30), holding that position until June as the number of COVID-SCs slowed and remained comparably low.

Thereafter, the Delta variant, characterized by being both more transmissible and more severe than both the ancestral strain or Alpha variant (31, 32), predominated through the start of SY 2021-22 until mid-December (21). During this period, FDA EUAs for COVID-19 vaccines were extended to those 12-15 years of age in May 2021 and finally to those 5-11 years of age in October 2021 (30), whereby all K-12 students, teachers, and staff were eligible for vaccination during the first half of SY 2021-22. Prior to the start of SY 2021-22, it was reported that almost 90% of teachers were vaccinated nationally (33). Expanded vaccine eligibility and high uptake among teachers and staff coincided with a return to in-person learning for nearly all schools in the US in SY 2021-22 (3, 5). In this SY, epidemiologic data peaks in January 2022 amid the domination of the Omicron variant, thus far characterized as the fastest spreading variant (34), were matched with the highest weekly counts of COVID-SCs. While we observed lower correlation of COVID-SCs with COVID-19 cases and percent PCR positivity in 2021-22 than in the previous year, correlations with new deaths and hospitalization rates were both higher. Strong correlation between COVID-SCs and hospitalization rates may suggest that COVID-SCs are not only associated with disease prevalence but also with the severity of the dominant circulating variant.

Various prevention measures were implemented in schools and districts to prevent the spread of SARS-CoV-2 (35), thereby, notionally reducing the number of COVID-SCs. However, disparities in their prevalence of implementation have been observed across locales and school poverty levels (35). Infection prevention measures include non-pharmaceutical interventions that can be rapidly implemented in schools, such as masking, social distancing, and quarantining, all of which have previously been shown to be effective at slowing influenza transmission in community congregate settings (36); at least one study documented that use of face masks reduces SARS-CoV-2 infection incidence in k-12 schools (37).

This study is subject to at least four limitations. First, reports are limited to publicly available data, and some closures may have been missed depending on how they were reported. Second, data may be incomplete for various reasons including, delays in identifying public announcements of school closures, incomplete or unavailable public announcements, or lags in data entry. Third, lengths of closure may be unknown when the specific date of reopening cannot be ascertained or because the school/district remains closed. And fourth, learning modality may not be specified in the data abstraction source (the announcement and/or website), and the data may therefore not capture all transitions from in-person to distance learning. However, these data were collected without burden to schools or districts and were readily available in near-real-time. These limitations likely lead to underestimation of the number and duration of school closures. Therefore, our results likely convey the lower range of the impact of COVID-SCs during this time period.

The COVID-SC data presented here were collected as part of an ongoing research project implemented to document how school closures occur outside of an influenza pandemic (1). In the absence of a true surveillance system, these were the best data available on COVID-SCs from the very early days of the pandemic. This project documented near-simultaneous nationwide closures implemented as a mitigation strategy during the spring of 2020 (2) as well as an unprecedented number of illness-related reactive school closures during the two subsequent school years, 2020-21 and 2021-22. These data could be used in conjunction with epidemiologic surveillance and other data for future computer simulations to explore the impact of the COVID-19 pandemic at various geographic levels and to help evaluate effectiveness of contemporaneous pandemic interventions. With SARS-CoV-2 /COVID-19 already established as an endemic disease and with the re-established circulation of other respiratory pathogens, local outbreaks of COVID-19, influenza, and other diseases will likely continue to occur, and some will cause reactive school closures. The continued monitoring of disease-related school closures should preserve the ability to detect their occurrence in near real-time as a component of community-based surveillance during pandemics and severe outbreaks. Additionally, ongoing surveillance for disease-related school closures would help understand their underlying causes, scale and distribution, and facilitate evaluation of their impact on schools and communities.

## Disclaimer

The opinions expressed by authors contributing to this journal do not necessarily reflect the opinions of the Centers for Disease Control and Prevention or the institutions with which the authors are affiliated.

## Data Availability

All data produced in the present study will be made available upon acceptance by a peer review journal.

## Acknowledgments

The authors would like to thank the data collection team, including our unit ORISE Fellows (Livvy Shafer, Pallavi Malla, and Larry Ayer) and contractors (Tamara Cummings, Jasminn Evans, Atea Francis, Elisha Gilbert, Esther Amoakohene, and Zaneta Oliver).

