## Supplemental Materials for "COVID-19-related school closures, United States, July 27, 2020 – June 30, 2022"

**Supplementary Material**

**Authors**

Nicole Zviedrite^1*^, Ferdous Jahan^1, 2^, Sarah Moreland^1,3^, Faruque Ahmed^1^, Amra Uzicanin^1^

**Affiliations**

^1^Centers for Disease Control and Prevention, Atlanta, Georgia, USA.

^2^Cherokee Nation Operational Solutions, LLC, Tulsa, Oklahoma, USA

^3^Oak Ridge Institute for Science and Education, Oak Ridge, Tennessee, United States of America

**Corresponding author***

Nicole Zviedrite, MPH, Community Interventions for Infection Control Unit, Division of Global Migration and Quarantine, National Center for Emerging Zoonotic Infectious Diseases, Centers for Disease Control and Prevention, 1600 Clifton Rd NE, MS V18-2, Atlanta, GA 30329,, 404-639-3961

### **ADDITIONAL DESCRIPTION OF METHODS**

To supplement the methods section, additional information is provided below. Included are methods to calculate the number of school days lost due to COVID-SCs, to conduct descriptive analyses of repeat closures for COVID-19, to describe the number and rate of COVID-SCs by state, and finally to conduct bivariate and multiple logistic regression analysis on school characteristics.

#### IN-PERSON SCHOOL DAYS LOST DUE TO COVID-SCS

In-person school days lost due to COVID-SCs were calculated by counting the total number of days between the date of closure (inclusive) and the date of reopening (exclusive), and then subtracting any planned closure days such as weekends, planned holidays, or teacher in-service days. The number of COVID-SCs by number of in-person school days lost were charted with whisker plots of the annual distributions of unplanned closure days overlain, using Power BI. Median number of in-person school days lost by state and by year were calculated and mapped using Power BI.

#### REPEAT CLOSURES FOR COVID-19

During the course of analysis, repeat closures of the same schools for COVID-19 were noted in the data within each school year. We describe the patterns seen across the study period.

#### COVID-SCS BY STATE

Cumulative incidence of COVID-SCs were calculated for the 2021/22 school year, with the number of COVID-SCs being the numerator and the total number of K-12 schools (public [1] & private [2], as reported by NCES) being the denominator. The number of COVID-SCs and the cumulative incidence of closures were mapped by state for the 2021/22 school year using Power BI.

#### BIVARIATE AND MULTIPLE LOGISTIC REGRESSION

Bivariate and multivariable logistic regression were performed using PROC LOGISTIC to examine both the unadjusted and adjusted odds ratios between COVID-SCs and certain school characteristics in the NCES data set for public schools (1), which comprise 98.4% of schools in our data set. Private schools accounted for the remaining 1.6%% of COVID-SCs and were excluded as the NCES Private School Survey data (2) did not fully include the corresponding variables found in public school data. The dependent variable was the number of unique schools closed for COVID-19 vs the total number of schools that did not close due to COVID-19, and the independent variables were urbanicity, student-teacher ratio, and percentage of students eligible for free or reduced-price school meals. Analysis was conducted using SAS 9.4 (SAS Institute Inc., Cary, North Carolina).

### **SUPPLEMENTARY RESULTS**

#### IN-PERSONSCHOOL DAYS LOST DUE TO COVID-19-RELATED SCHOOL CLOSURES

See Figures S1 and S2.

##### **Figure S1. Number of In-Person School Days Lost Due to * COVID-19-related School Closures† per Closure by Closure Type‡§, United States, July 27, 2020 – June 30, 2022**

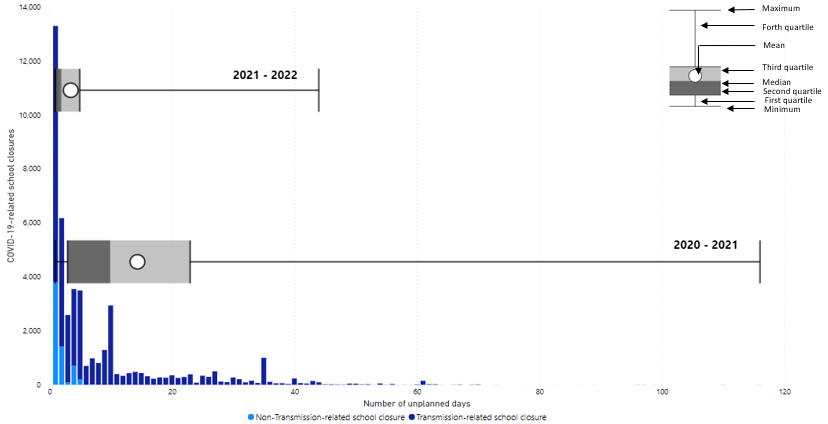

*In-Person school days lost includes only unplanned closure days and excludes weekends and planned closure days (holidays, teacher workdays, etc.). These are verified using school or school district calendars when available. Academic year 2020 – 2021: median 10, mean 14, range 1-116. Academic year 2021 – 2022: median 2, mean 3, range 1-44.

^†^School closure is defined as a transition from being open to being closed for in-person instruction excluding any scheduled days off; fully in-person and hybrid learning modalities are classified as open, and fully remote and closed are classified as closed.

^‡^Transmission-related reasons include COVID-19 cases, suspected cases, increased student absenteeism, increased staff absenteeism, cluster or widespread transmission in the community, state or local guidance/mandate to close schools in response to COVID-19, to clean/disinfect school facilities, and other.

^§^Non-transmission-related reasons include COVID-19 vaccinations and side effects of vaccination of staff/students, teacher/staff shortage, for student/staff mental health, and other reasons associated with COVID-19.

##### **Figure S2. Median number of in-person school days lost due to COVID-19-related school closure^*†‡^**^§^ **by state, United States, July 27, 2020 - June 30, 2022**

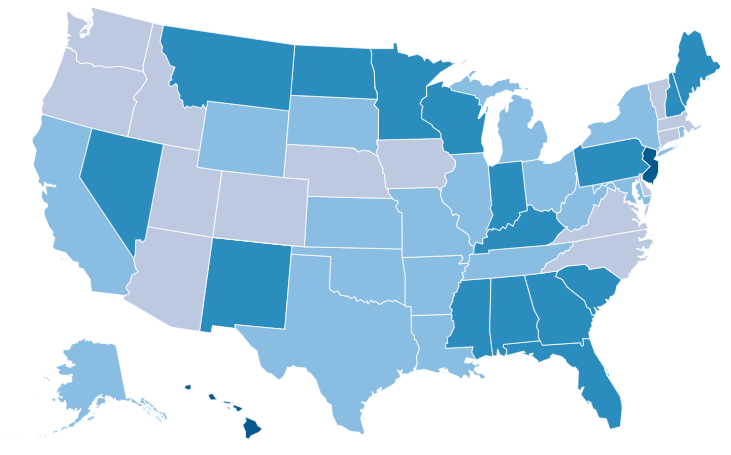

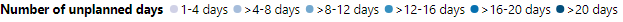

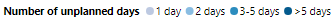

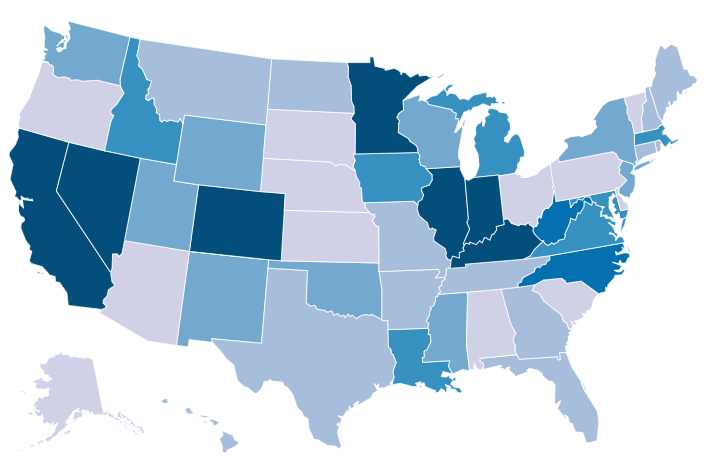

**2020 – 2021 school year**

**2021 – 2022 school year**

^*^School closure is defined as a transition from being open to being closed for in-person instruction excluding any scheduled days off; fully in-person and hybrid learning modalities are classified as open, and fully remote and closed are classified as closed.

^†^In-person school days lost includes only unplanned closure days and excludes weekends and planned closure days (holidays, teacher workdays, etc.). These are verified using school or school district calendars when available.

^‡^Transmission-related reasons include COVID-19 cases, suspected cases, increased student absenteeism, increased staff absenteeism, cluster or widespread transmission in the community, state or local guidance/mandate to close schools in response to COVID-19, to clean/disinfect school facilities, and other.

^§^Non-transmission-related reasons include COVID-19 vaccinations and side effects of vaccination of staff/students, teacher/staff shortage, for student/staff mental health, and other reasons associated with COVID-19.

#### REPEAT CLOSURS FOR COVID-19

Repeat closures for COVID-19 were concentrated in HHS 4 for both 2020-2021 and 2021-2022 school year.(Table S1). For 2020-2021 school year, more than one-third of the COVID-19-related repeat closures were observed in suburban areas, while majority of the repeat closure for 2021-2022 school year were concentrated in city areas.

See Table S1.

##### **Table S1. Recurrence of COVID-related school closures among unique schools*– United States, July 27, 2020 – June 30, 2022**

| **Characteristics of schools closed** | Unique schools,  n (%) | Unique schools by number of closures, n (n%) | | | | |
| --- | --- | --- | --- | --- | --- | --- |
|  |  | Single closure occurrence, n (%) | Multiple closures, n (%) | | | |
|  |  |  | Total (≥ 2X)^‡^ | 2X | 3-4X | ≥ 5X^‡^ |
| **2020-2021 School Year** |  |  |  |  |  |  |
| Total^§^ | 16,890 | 14,854 (87.9) | 2,036 (12.1) | 1,756 (10.4) | 267 (1.6) | 13 (0.1) |
| Urbanicity |  |  |  |  |  |  |
| City | 5,997 (35.5) | 5,351 (36.0) | 646 (31.7) | 570 (32.5) | 73 (27.3) | 3 (23.1) |
| Suburban | 5,297 (31.4) | 4,585 (30.9) | 712 (35.0) | 633 (36.1) | 72 (27.0) | 7 (53.9) |
| Town | 1,687 (10.0) | 1,469 (9.9) | 218 (10.7) | 174 (9.9) | 43 (16.1) | 1 (7.7) |
| Rural | 3,630 (21.5) | 3,174 (21.4) | 456 (22.4) | 375 (21.4) | 79 (29.6) | 2 (15.4) |
| Not specified | 279 (1.7) | 275 (1.9) | 4 (0.2) | 4 (0.2) | 0 | 0 |
| HHS Region^¶^ |  |  |  |  |  |  |
| HHS 1 | 1,049 (6.2) | 851 (5.7) | 198 (9.7) | 154 (8.8) | 42 (15.7) | 2 (15.4) |
| HHS 2 | 2,795 (16.0) | 2,319 (15.6) | 386 (19.0) | 341 (19.4) | 44 (16.5) | 1 (7.7) |
| HHS 3 | 2,399 (14.2) | 2,019 (13.6) | 380 (18.7) | 298 (17.0) | 76 (28.5) | 6 (46.2) |
| HHS 4 | 3,455 (20.5) | 2,997 (20.2) | 458 (22.5) | 407 (23.2) | 48 (18.0) | 3 (23.1) |
| HHS 5 | 3,254 (19.3) | 2,910 (19.6) | 344 (16.9) | 317 (18.1) | 27 (10.1) | 0 |
| HHS 6 | 926 (5.5) | 821 (5.5) | 105 (5.2) | 87 (5.0) | 18 (6.7) | 0 |
| HHS 7 | 566 (3.4) | 544 (3.7) | 22 (1.1) | 22 (1.3) | 0 | 0 |
| HHS 8 | 782 (4.6) | 694 (4.7) | 88 (4.3) | 86 (4.9) | 2 (0.8) | 0 |
| HHS 9 | 1,418 (8.4) | 1,384 (9.3) | 34 (1.7) | 33 (1.9) | 1 (0.4) | 0 |
| HHS 10 | 336 (2.0) | 315 (2.1) | 21 (1.0) | 11 (0.6) | 9 (3.4) | 1 (7.7) |
| **2021-2022 School Year** |  |  |  |  |  |  |
| Total^§^ | 19,871 | 15,404 (77.5) | 4,467 (22.5) | 3,620 (18.2) | 686 (3.5) | 161 (0.8) |
| Urbanicity |  |  |  |  |  |  |
| City | 7,338 (36.9) | 4,653 (30.2) | 2,685 (60.1) | 2,173 (60.0) | 430 (62.7) | 82 (50.9) |
| Suburban | 5,966 (30.0) | 4,948 (32.1) | 1,018 (22.8) | 753 (20.8) | 201 (29.3) | 64 (39.8) |
| Town | 2,215 (11.2) | 1,996 (13.0) | 219 (4.9) | 204 (5.6) | 9 (1.3) | 6 (3.7) |
| Rural | 4,286 (21.6) | 3,747 (24.3) | 539 (12.1) | 484 (13.4) | 46 (6.7) | 9 (5.6) |
| Not specified | 66 (0.3) | 60 (0.4) | 6 (0.0) | 6 (0.2) | 0 | 0 |
| HHS Region^¶^ |  |  |  |  |  |  |
| HHS 1 | 554 (2.8) | 517 (3.4) | 37 (0.8) | 27 (0.8) | 10 (1.5) | 0 |
| HHS 2 | 1,334 (6.7) | 1,200 (7.8) | 134 (3.0) | 131 (3.6) | 3 (0.4) | 0 |
| HHS 3 | 2,029 (10.2) | 1,534 (10.0) | 495 (11.1) | 338 (9.3) | 152 (22.2) | 5 (3.1) |
| HHS 4 | 4,726 (23.8) | 4,084 (26.5) | 642 (14.4) | 591 (15.3) | 51 (7.4) | 0 |
| HHS 5 | 3,946 (19.9) | 2,363 (15.3) | 1,583 (35.4) | 1,322 (36.5) | 227 (33.1) | 34 (21.1) |
| HHS 6 | 3,362 (16.9) | 2,873 (18.7) | 489 (11.0) | 380 (10.5) | 94 (13.7) | 15 (9.3) |
| HHS 7 | 1,206 (6.1) | 918 (6.0) | 288 (6.5) | 202 (5.6) | 68 (9.9) | 18 (11.2) |
| HHS 8 | 930 (4.7) | 482 (3.1) | 448 (10.0) | 381 (10.5) | 9 (1.3) | 58 (36.0) |
| HHS 9 | 768 (3.9) | 727 (4.7) | 41 (0.9) | 23 (0.6) | 0 | 18 (11.2) |
| HHS 10 | 1,016 (5.1) | 706 (4.6) | 310 (6.9) | 225 (6.2) | 72 (10.5) | 13 (8.1) |

^*^Unique school: each school experiencing closure was counted only once.

^†^School year: 2020-2021 (July 27, 2020 to June 30, 2021), 2021-2022 (Aug 1, 2021 to Jun 30, 2022).

^‡^The maximum number of repeat closures was seven times during 2020-2021 school year and eight times during 2021-2022 school year.

^§^Total row presented with row percent, all else reported with column percent.

^¶^Regions of the United States Department of Health & Human Services (HHS) [3].

#### COVID-SCS BY STATE

See Figure S3.

##### **Figure S3.** **Number of COVID-19-related school closures^*†‡^ and cumulative incidence of closures^§^ by states, United States, August 1, 2021 – June 30, 2022**

Fig. S3.b

Fig. S3.a

^*^School closure is defined as a transition from being open to being closed for in-person instruction excluding any scheduled days off; fully in-person and hybrid learning modalities are classified as open, and fully remote and closed are classified as closed.

^†^Transmission-related reasons include COVID-19 cases, suspected cases, increased student absenteeism, increased staff absenteeism, cluster or widespread transmission in the community, state or local guidance/mandate to close schools in response to COVID-19, to clean/disinfect school facilities, and other.

^‡^Non-transmission-related reasons include COVID-19 vaccinations and side effects of vaccination of staff/students, teacher/staff shortage, for student/staff mental health, and other reasons associated with COVID-19.

**^§^**The denominator was the total number of k-12 schools (public &private) in 2021 NCES databases [1,2].

#### REGRESSION ANALYSIS

During the 2020-2021 school year, schools located in rural areas (aOR 0·47, 95% CI 0·45-0·50), towns (aOR 0·54, 95% CI 0·51-0·58), or suburban areas (aOR 0·67, 95% CI 0·64-0·71) (Table S2) had lower odds of experiencing closure than those located in cities. Meanwhile, the chance of school closure was significantly lower for the highest quartile of student-teacher ratio (aOR 0·72, 95% CI 0·68-0·76). Schools in the upper two quartiles of students eligible for free or reduced lunch showed significant lower odds of COVID-SCs as compared to first quartile (Q3: aOR 0·87, 95% CI 0·82-0·92, and Q4: aOR 0·89, 95% CI 0·84-0·94).

Similar to the previous SY, during the 2021-2022 SY, schools located in rural areas (aOR 0·54, 95% CI 0·52-0·57), towns (aOR 0·65, 95% CI 0·61-0·69), or suburban areas (aOR 0·69, 95% CI 0·66-0·72) had lower odds of experiencing a COVID-SC than those located in cities (Table S2). Schools in the 3^rd^ quartile of student-teacher ratio showed higher odds of closure (aOR 1·25, 95% CI 1·18-1·31) followed by the 2^nd^ quartile (aOR 1·18, 95% CI 1·12-1·24) when compared to the lowest quartile, while schools in the highest quartile of student-teacher ratio showed lower odds (aOR 0·77, 95% CI 0·73, 0·814). The chance of school closures increased as the percentage of students eligible for free or reduced lunch increased, the highest odds of closures were observed in the highest quartile (aOR 1·61, 95% CI 1·53, 1·70) when compared to the lowest quartile (Table S2).

See Table S2.

##### **Table S2. Selected characteristics of COVID-19-related unique* public school closures by school year^†^ — United States, July 27, 2020—June 30, 2022**^‡^

|  | **2020–2021**^§^ | | | | **2021–2022**^§^ | | | |
| --- | --- | --- | --- | --- | --- | --- | --- | --- |
|  | **Unadjusted OR (95% CI)** | **P** | **Adjusted OR (95% CI)** | **P** | **Unadjusted OR (95% CI)** | **P** | **Adjusted OR (95% CI)** | **P** |
| **Urbanicity** |  |  |  |  |  |  |  |  |
| City | Ref. |  | Ref. |  | Ref. |  |  |  |
| Rural | 0.54 (0.51, 0.56) | <0.001 | 0.47 (0.45, 0.50) | <0.001 | 0.49 (0.47, 0.51) | <0.001 | 0.54 (0.52, 0.57) | <0.001 |
| Town | 0.55 (0.52, 0.59) | <0.001 | 0.54 (0.51, 0.58) | <0.001 | 0.57 (0.54, 0.60) | <0.001 | 0.65 (0.61, 0.69) | <0.001 |
| Suburban | 0.72 (0.69, 0.75) | <0.001 | 0.67 (0.64, 0.71) | <0.001 | 0.65 (0.62, 0.67) | <0.001 | 0.69 (0.66, 0.72) | <0.001 |
| **Student-teacher ratio**^¶^ |  |  |  |  |  |  |  |  |
| <=Q1 | Ref. |  | Ref. |  | Ref. |  | Ref. |  |
| >Q1-Q2 | 1.13 (1.07, 1.19) | <0.001 | 1.06 (1.00, 1.12) | 0.040 | 1.26 (1.21, 1.32) | <0.001 | 1.18 (1.12, 1.24) | <0.001 |
| >Q2-Q3 | 1.06 (1.01, 1.11) | <0.001 | 0.95 (0.90, 1.00) | 0.064 | 1.35 (1.29, 1.42) | <0.001 | 1.25 (1.18, 1.31) | <0.001 |
| >Q3 | 0.83 (0.79, 0.87) | <0.001 | 0.72 (0.68, 0.76) | <0.001 | 0.91 (0.87, 0.95) | <0.001 | 0.77 (0.73, 0.81) | <0.001 |
| **Percent of students eligible for free or reduced school meals**^#**^ |  |  |  |  |  |  |  |  |
| <=Q1 | Ref. |  | Ref. |  | Ref. |  | Ref. |  |
| >Q1-Q2 | 0.91 (0.86, 0.96) | 0.003 | 0.96 (0.91, 1.02) | 0.156 | 1.30 (1.24, 1.37) | <0.001 | 1.29 (1.23, 1.35) | <0.001 |
| >Q2-Q3 | 0.90 (0.86, 0.95) | <0.001 | 0.87 (0.82, 0.92) | <0.001 | 1.60 (1.53, 1.68) | <0.001 | 1.47 (1.39, 1.54) | <0.001 |
| >Q3 | 1.02 (0.97, 1.08) | <0.001 | 0.89 (0.84, 0.94) | <0.001 | 2.05 (1.95, 2.16) | <0.001 | 1.61 (1.53, 1.70) | <0.001 |

^*^Unique school: each school experiencing closure was counted only once.

**^†^**School year: 2020-2021 (July 27, 2020 to June 30, 2021), 2021-2022 (Aug 1, 2021 to Jun 30, 2022).

^‡^Bivariate and multivariate logistic regression were used for the unadjusted and adjusted odds ratios respectively. Dependent variable: The total number of unique schools closed during the period vs the total number of schools that didn’t close. Independent variables: urbanicity, student-teacher ratio, and percentage of students eligible for free or reduced-price school meals.

^§^The total number of unique public schools that matched with the NCES public school data [1] and used for this analysis was 16,188 for 2020-2021 school year and 19,763 for 2021-2022 school year.

^¶^Student-teacher ratio by year: 2020-2021 - The lower quartile (Q1), median (Q2), and upper quartile (Q3) was 12.05, 14,27, and 16.84 respectively. 2021-2022 - The lower quartile (Q1), median (Q2), and upper quartile (Q3) was 12.37, 14,54, and 16.87 respectively.

^#^Percent of students eligible for free or reduced meals by year: 2020-2021 - The lower quartile (Q1), median (Q2), and upper quartile (Q3) was 32.11, 54.16, and 80.76 respectively. 2021-2022 - The lower quartile (Q1), median (Q2), and upper quartile (Q3) was 36.83, 59.97, and 84.80 respectively.

^**^Private schools were excluded from this analysis; the data are available for public schools only [1].
